# Development of a Natural Language Processing Algorithm for the Classification of Suspicious Liver Lesions from Radiology Reports

**DOI:** 10.1101/2021.12.15.21267875

**Authors:** Jacob Johnson, Kaneel Senevirathne, Lawrence Ngo

## Abstract

Here, we developed and validated a highly generalizable natural language processing algorithm based on deep-learning. Our algorithm was trained and tested on a highly diverse dataset from over 2,000 hospital sites and 500 radiologists. The resulting algorithm achieved an AUROC of 0.96 for the presence or absence of liver lesions while achieving a specificity of 0.99 and a sensitivity of 0.6.

## Introduction

Liver lesions are focal abnormalities in the organ’s parenchyma, which can often be visualized using various forms of medical imaging. While most lesions are benign, a significant subset can be cancerous. Computer tomography (CT) medical imaging exams are the most common method of identifying liver lesions. While CT exams are reliable and widely used, there are challenges in diagnosis due to technical factors (e.g. motion artifact and contrast bolus timing) and non-technical factors (e.g. human error), and these can decrease the confidence of diagnosis. Similarly, the language in radiology reports can vary and significantly increase the complexity associated with developing algorithms for classification of liver lesions (1,2).

Previous work on algorithm development for radiologist reports used lexical rules to classify medical imaging reports (3,4). While these algorithms generated moderate performance, such approaches had difficulty dealing with different classes of diagnosis. Furthermore, it is very labor intensive to develop and may not generalize across different languages and pathologies. Recent work has used more sophisticated strategies such as convolutional neural networks and achieved significantly higher levels of performance in diagnosis in pathologies such as pulmonary embolism (5,6).

However, generalizability of these algorithms remains a question. Most of the previous studies have used datasets collected from the same or a few number of hospital sites (3,5,7). Thus, these datasets could be biased towards their regional idiosyncrasies in dictation. If such algorithms are tested in different hospital settings, weaknesses in performance may arise due to the homogeneity of the dataset utilized in the training phase.

In our current study, we developed a highly generalizable NLP algorithm to detect suspicious liver lesions from radiologist reports. Our training, validation and testing datasets were collected from a very diverse distribution. Our datasets were from numerous hospital sites spread across the United States. Due to the variety of the hospital sites our datasets represent diverse radiology reports and CT protocols including a variety of lexical rules used in different regions. The high level of generalizability of our algorithm increases the chances by which such an algorithm could be deployed in any given hospital site without significant degradation in performance.

## Methods

This project was performed with an IRB approval for waiver of consent by an external IRB, Western IRB. Radiology reports for this study were obtained from Virtual Radiologic (vRad) and were de-identified before use. The reports were sourced from over 2,000 hospital sites and 500 radiologists across the United States. An initial set of radiology reports were reviewed by LN, a radiologist with 5 years of radiology experience and 13 years of general medical experience with the help of a NLP tool developed in-house based on lexical rules to categorize reports as either “liver lesion present” or “liver lesion absent.” This tool was used to mine a database of over 200,000 patient reports to identify reports of interest to manually label, which was performed by a group of medical professionals with at least 8 years of experience in the medical field.

We used a deep learning bidirectional transformer (hereafter called Liver-BERT) as our liver lesion classification algorithm (8). For training the model, a total of 178,245 radiologist reports were collected. The training set consisted of 15,454 positive exams and 162,791 negative exams. The validation set consisted of 1,701 positive exams and 18,105 negative exams. No preprocessing was performed on any of the reports prior to training. After preparing data we used Liver-BERT with pre-trained weights and further trained using our training dataset which resulted in an accuracy of 0.97 on the validation set. We additionally tested the model on a hold-out test-set of 3,568 reports that were labeled by the same team of medical professionals as above.

We then evaluated the performance of Liver-BERT on the test set by measuring the sensitivity, specificity, positive predictive value (PPV) and the negative predictive value (NPV).

We also measured the receiver operating characteristic (ROC) curve and the area under the ROC curve (AUROC) to assess the model further.

## Results

The incidence of liver lesions within the test set was 0.03 (117 positive and 3,451 negative). Liver-BERT achieved a final AUROC of 0.96. The model sensitivity and specificity was 0.60 and 0.99, respectively. The summary of our model performance on liver lesion reports is shown in Table 1.

**Table 1:** Performance summary of Liver-BERT algorithm

|  | Incidental Liver Lesion |
| --- | --- |
| AUROC | 0.96 |
| Sensitivity | 0.60 |
| Specificity | 0.99 |
| PPV | 0.58 |
| NPV | 0.99 |

## Discussion

In this study, we developed a deep-learning natural language processing algorithm for classification of liver lesions on radiology reports. We used Liver-BERT, a bidirectional transformer model with pre-trained weights and trained it further on a highly diverse dataset of liver lesion reports. Several components of our approach are novel and may contribute to the high performance of this algorithm. Initially, we used a lexicon rules based algorithm to assist labeling the liver lesion reports of the training dataset. Due to the large sample size of our training set, to supplement this methodology, we further employed an image based liver lesion model. This helped us to identify discrepancies between the image generated and lexicon rules based algorithm generated labels and adjust incorrect labels of the training set accordingly.

Furthermore, several aspects of our study demonstrate that our Liver-BERT based liver lesion classification model is highly generalizable. First, in our study we did not include a data preprocessing step before training and inference. However, in previous approaches based on deep learning in medical report classification, only impressions were included in the training dataset. Such an approach may be susceptible to differences in report formatting, language and report generation style in various hospital sites. For instance, some hospitals prefer listing their impressions at the very beginning of the report while some sites prefer listing at the bottom. Furthermore, some radiologists may label their impression with alternative names. (e.g. conclusion, opinion, decision). Thus, by including the entire report without any preprocessing, we were able to ensure that our model is robust across such variations in formatting, language and style.

Our dataset was collected from over 2,000 different hospital sites and represented over 500 different radiologists. This makes our dataset highly distributed, thus increasing the generalizability of our algorithm. Such a level of generalizability is crucial if the final outcome is to deploy the model to different hospital sites. Additionally, given the large size of our test set, we were able to stratify the performance of the algorithm to fit into any clinical setting.

Finally, to our understanding, our current dataset of liver lesion reports represents the largest dataset used for the purpose of using NLP for classification for suspicious liver lesions. This ensures not only that our performance is high, but also that the model is robust to heterogeneity in reports that may come from a variety of sources.

## Data Availability

The data from this study is not publicly available.

## Acknowledgements

We would like to thank Nadine Ly for her assistance in paper draft editing, Edin Mesanovic and Nina Perez for providing a subset of radiology annotations, and Brian Baker, Christine Lamoureux, Ian Driscoll, and Jerry Lohr from Virtual Radiologic for their support with clinical data.

## Supplemental

NLP criteria for a suspicious liver lesion:

- Positive if the report mentions:
  - Imaging is recommended unequivocally to further evaluate a liver lesion
  - Report mentions a cystic mass.
  - Liver lesion that is indeterminate with or without follow up recommendation.
  - Suspected hemangioma but recommend follow up
  - Something in findings related to a suspicious liver lesion but never recommends anything for it. Report does not call it indeterminate.
  - Known suspicious liver lesions that may or may not be described in more detail.
- Negative if the report mentions:
  - Cyst, too small to characterize.
  - Lesion favored to be a cyst.
  - Stable hypodensity with no other description.
  - Small hypodensity not otherwise characterized.
  - Liver granulomas.
  - Lipoma.

